# Diabetes and Mortality Among 1.6 Million Adult Patients Screened for SARS-CoV-2 in Mexico

**DOI:** 10.1101/2020.11.25.20238345

**Authors:** Orison O. Woolcott, Juan P. Castilla-Bancayán

## Abstract

**Background:** Whether diabetes is associated with COVID-19-related mortality remains unclear.

**Methods:** In this retrospective case-series study we examined the risk of death associated with self-reported diabetes in symptomatic adult patients with laboratory-confirmed COVID-19 who were identified through the System of Epidemiological Surveillance of Viral Respiratory Disease in Mexico from January 1 through November 4, 2020. Survival time was right-censored at 28 days of follow-up.

**Results:** Among 757,210 patients with COVID-19 included in the study, 120,476 (16%) had diabetes and 80,616 died. Patients with diabetes had a 49% higher relative risk of death than those without diabetes (Cox proportional-hazard ratio; 1.49 (95% confidence interval [CI], 1.47-1.52), adjusting for age, sex, smoking habit, obesity, hypertension, immunodeficiency, and cardiovascular, pulmonary, and chronic renal disease. The relative risk of death associated with diabetes decreased with age (P=0.004). The hazard ratios were 1.66 (1.58-1.74) in outpatients and 1.14 (1.12-1.16) in hospitalized patients. The 28-day survival for inpatients with and without diabetes was, respectively, 73.5% and 85.2% for patients 20-39 years of age; 66.6% and 75.9% for patients 40-49 years of age; 59.4% and 66.5% for patients 50-59 years of age; 50.1% and 54.6% for patients 60-69 years of age; 42.7% and 44.6% for patients 70-79 years of age; and 38.4% and 39.0% for patients 80 years of age or older. In patients without COVID-19 (878,840), the adjusted hazard ratio for mortality was 1.78 (1.73-1.84).

**Conclusion:** In symptomatic adult patients with COVID-19 in Mexico, diabetes was associated with higher mortality. This association decreased with age.

## INTRODUCTION

As of November 15, 2020, over 53.7 million people worldwide have been infected with SARS-CoV-2, the virus that causes COVID-19. Nearly 1.3 million people have died due to COVID-19.^1^

Patients with COVID-19 who have diabetes are at increased risk of hospitalization,^2,3^ admission to intensive care unit,^4^ and intubation,^3,5^ compared with those without diabetes. Although diabetes is common in fatal cases of COVID-19,^6-10^ whether diabetes is associated with COVID-19-related mortality remains unclear. Some studies have shown an association between diabetes and mortality in subjects with COVID-19,^4,6,7,9-14^ but others have not confirmed this relationship.^2,3,8,15-18^ These differences could be related to under-adjustment,^9,12^ the relatively small number of subjects to estimate mortality risk,^4,8,9,14,16,18^ the use of composite outcomes,^2,12^ the analysis of severe COVID-19 cases or critically ill patients only,^9,10,15,17^ the age of study participants and lack of stratification by age,^3,6,7,10-12^ and the inclusion of unconfirmed cases.^6,13^

Given the association of diabetes with severe COVID-19, specific guidelines for the treatment of patients with COVID-19 and diabetes have been proposed.^19,20^ However, the association of diabetes with mortality across age groups in outpatients and hospitalized patients has not been well studied. Thus, clarification on this aspect may have clinical implications for risk stratification. The aim of the present study was to examine the risk of death associated with diabetes in symptomatic adult patients with COVID-19 confirmed by laboratory.

## METHODS

### Study design and population

We conducted a retrospective case-series study using data from a large convenience sample of symptomatic patients with viral respiratory disease who were screened for SARS-CoV-2 using real-time reverse-transcriptase–polymerase-chain-reaction assay on samples obtained through oropharyngeal or nasopharyngeal swabs.^21^ Patients were identified through the System of Epidemiological Surveillance of Viral Respiratory Disease in Mexico from January 1 through November 4, 2020. We included patients who were admitted from January 1 through October 7, 2020 in such a way that each patient had a 28-day follow-up unless the event (death) occurred first. We excluded patients younger than 20 years of age or coded as pregnant, and those who did not have test results for SARS-CoV-2 (Figure S1 in the Supplementary Appendix).

Since this study involved the analysis of publicly available de-identified data only, institutional-review-board review was not required, as outlined in the Federal Policy for the Protection of Human Subjects (detailed in 45 CFR part 46).^22^

### Study setting

Mexico has an estimated population of 127.6 million.^23^ Adults 65 years of age or older represent 6.2% of the population.^24^ In Mexico, the System of Epidemiological Surveillance of Viral Respiratory Disease keeps track of suspected cases of viral respiratory disease, including COVID-19 cases, through reports from 475 Viral Respiratory Disease Monitoring Units (USMER) and all healthcare centers (non-USMER) located nationwide. The USMER reports all suspected cases with severe respiratory symptoms but only 10% of all suspected cases with mild symptoms. The non-USMER reports all cases of severe acute respiratory infection.^21^

### Data source

Data at the individual level were obtained from the publicly available COVID-19 online dataset updated daily by the Secretary of Health of Mexico (https://www.gob.mx/salud/documentos/datos-abiertos-bases-historicas-direccion-general-de-epidemiologia). All data on demographic and pre-existing comorbidities were obtained through a standardized questionnaire.^21^

### Definitions

According to the guidelines from the Secretary of Health of Mexico, a suspected case of viral respiratory disease was defined as a subject who presented, in the last 10 days, cough, dyspnea, fever, or headache, and at least one of the following signs or symptoms: myalgias, arthralgias, odynophagia, chills, chest pain, rhinorrhea, polypnea, anosmia, dysgeusia, or conjunctivitis.^21^ These guidelines, released on August 2020, are an update of the guidelines in which a suspected case of viral respiratory disease was defined as a subject who presented, in the last 7 days, cough, fever, or headache, accompanied with at least one of the following signs or symptoms: dyspnea, myalgias, arthralgias, odynophagia/sore throat, rhinorrhea, conjunctivitis, or chest pain.^25^ In the present study, a COVID-19 case was defined as a patient with suspected viral respiratory disease who had SARS-CoV-2 infection confirmed by reverse-transcriptase–polymerase-chain-reaction test. Patients who tested negative for SARS-CoV-2 were referred to as non-COVID-19 cases, regardless of epidemiological association with COVID-19.

### Statistical analyses

Incidence rates of death were expressed as cases per 100,000 person-days. We used Cox proportional-hazards regression to calculate the hazard ratio and 95% confidence intervals (CIs) for mortality. There was no violation of proportional-hazards assumption. Survival time was right-censored at 28 days of follow-up from the admission date (date of the patient’s visit). Multivariate analyses included adjustment for age, sex, smoking habit, obesity, hypertension, cardiovascular disease, chronic obstructive pulmonary disease, asthma, chronic kidney disease, and immunodeficiency. These variables were chosen based on our judgment as they have been associated with the severity of COVID-19 or mortality.^13,26,27^ Cox regression models were adjusted for age using a five-knot restricted cubic spline fitting for age.^28^ Analyses within each age group (20 to 39, 40 to 49, 50 to 59, 60 to 69, 70 to79, and ≥80 years) were adjusted for age as a continuous variable. We tested for interactions between diabetes and age, diabetes and sex, and diabetes and type of patient care (outpatient vs. inpatient). The trends for the hazard ratios across age groups were tested using weighted linear regression. The probability weights were obtained from the inverse of the variance of the risk estimates. Since missing data among predictors included in our regression models represented less than 0.75%, missing data were not imputed. A complete-case analysis was performed. There were no missing data on age, sex, date of hospital admission, date of symptoms onset, or date of death.

We conducted three sensitivity analyses to assess the robustness of our findings: 1) full models with further adjustment for pneumonia, admission to intensive care unit, intubation, and time from symptoms onset to admission; 2) multilevel mixed-effect survival regression models to assess the possible effect of geographical differences on our risk estimates;^29^ and 3) comparison of hazard ratios from analysis restricted to cases admitted before and after August 1 to address the possible influence of changes to the definition of suspected viral respiratory disease.^21,25^ We conducted stratified analysis according to age and sex, in outpatients and inpatients. We used the log-rank test to compare survival curves. All p values were two-sided. All analyses were performed using Stata 14 (StataCorp LP, TX).

## RESULTS

From January 1 through November 4, 2020, 2,445,709 symptomatic patients with viral respiratory disease of all ages were reported. In total, 1,650,432 patients met the study inclusion criteria. We excluded 197 patients who had implausible admission dates relative to their date of death. We also excluded 14,185 patients who had missing data (0.86%) on diabetes, smoking habit, obesity, hypertension, cardiovascular disease, chronic obstructive pulmonary disease, asthma, chronic kidney disease, immunodeficiency, pneumonia, intubation, and admission to intensive care unit (Table S1). Main analysis involved 757,210 adult patients with laboratory-confirmed COVID-19. We also included 878,840 adult patients who tested negative for SARS-CoV-2. In patients with COVID-19, the median age was 44 years (IQR, 33-56); 120,476 (15.9%) had diabetes (Table 1). The proportion of patients with diabetes was 10.4% among outpatients (575,866) and 33.3% among inpatients (181,344).

**Table 1.**
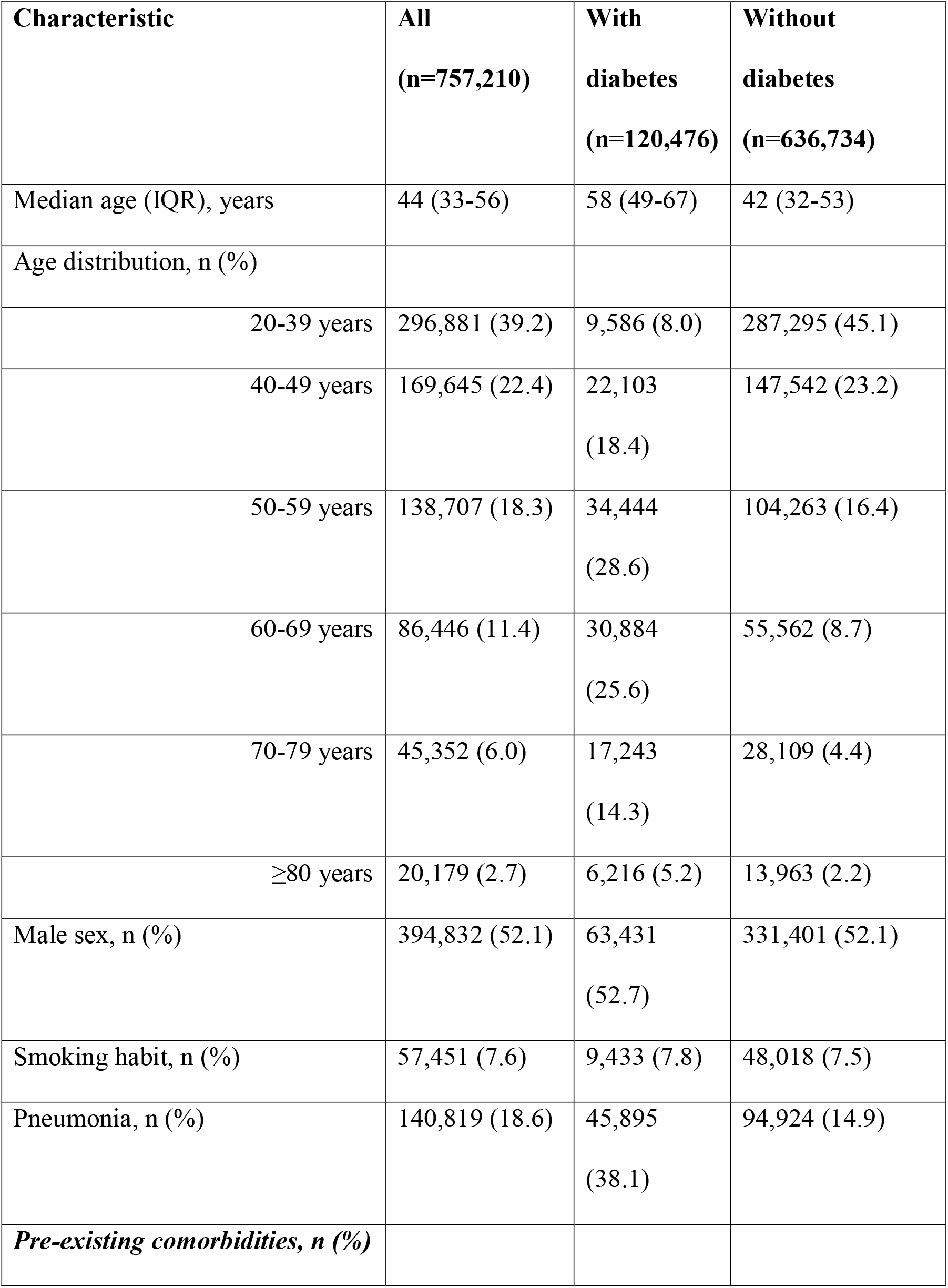
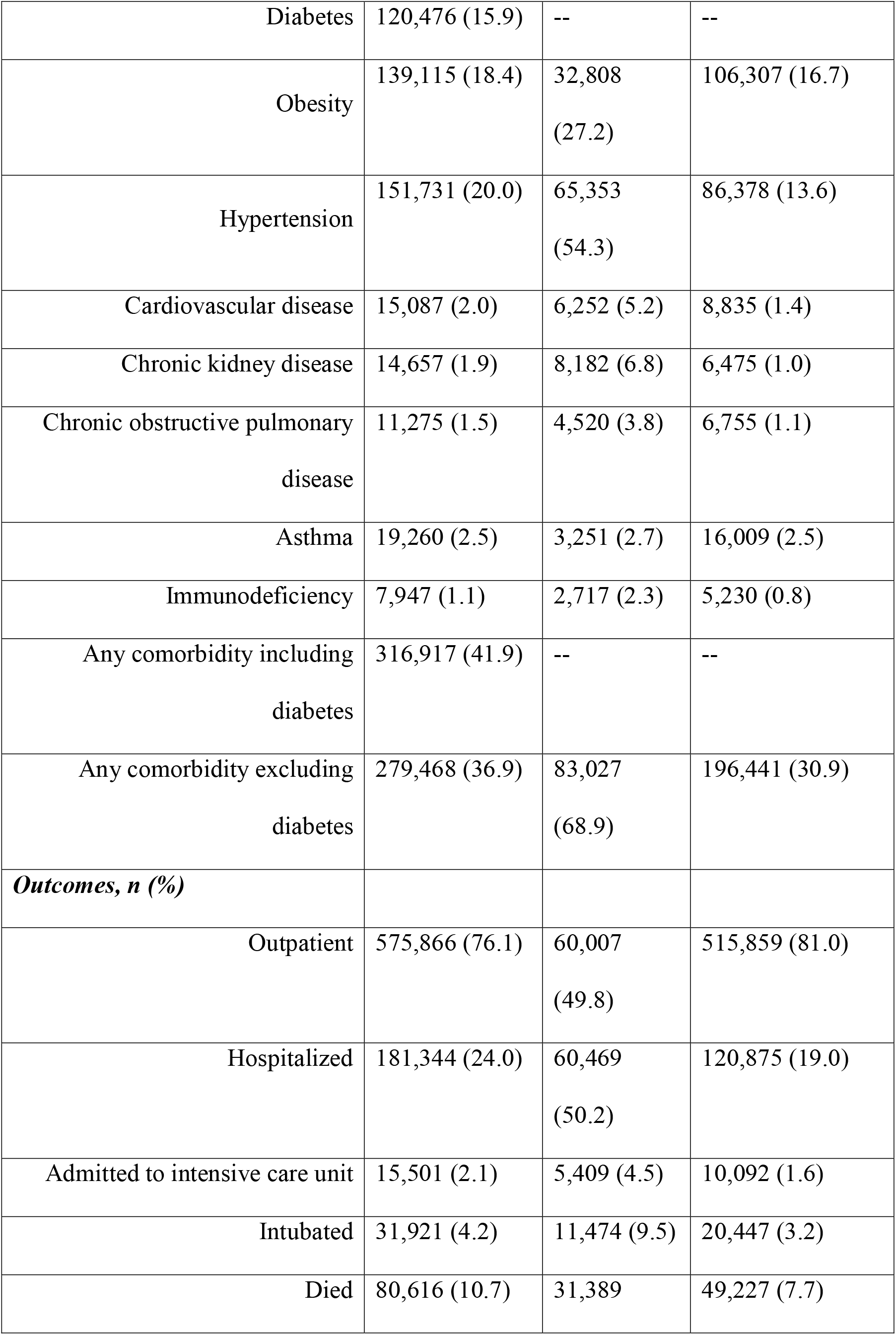
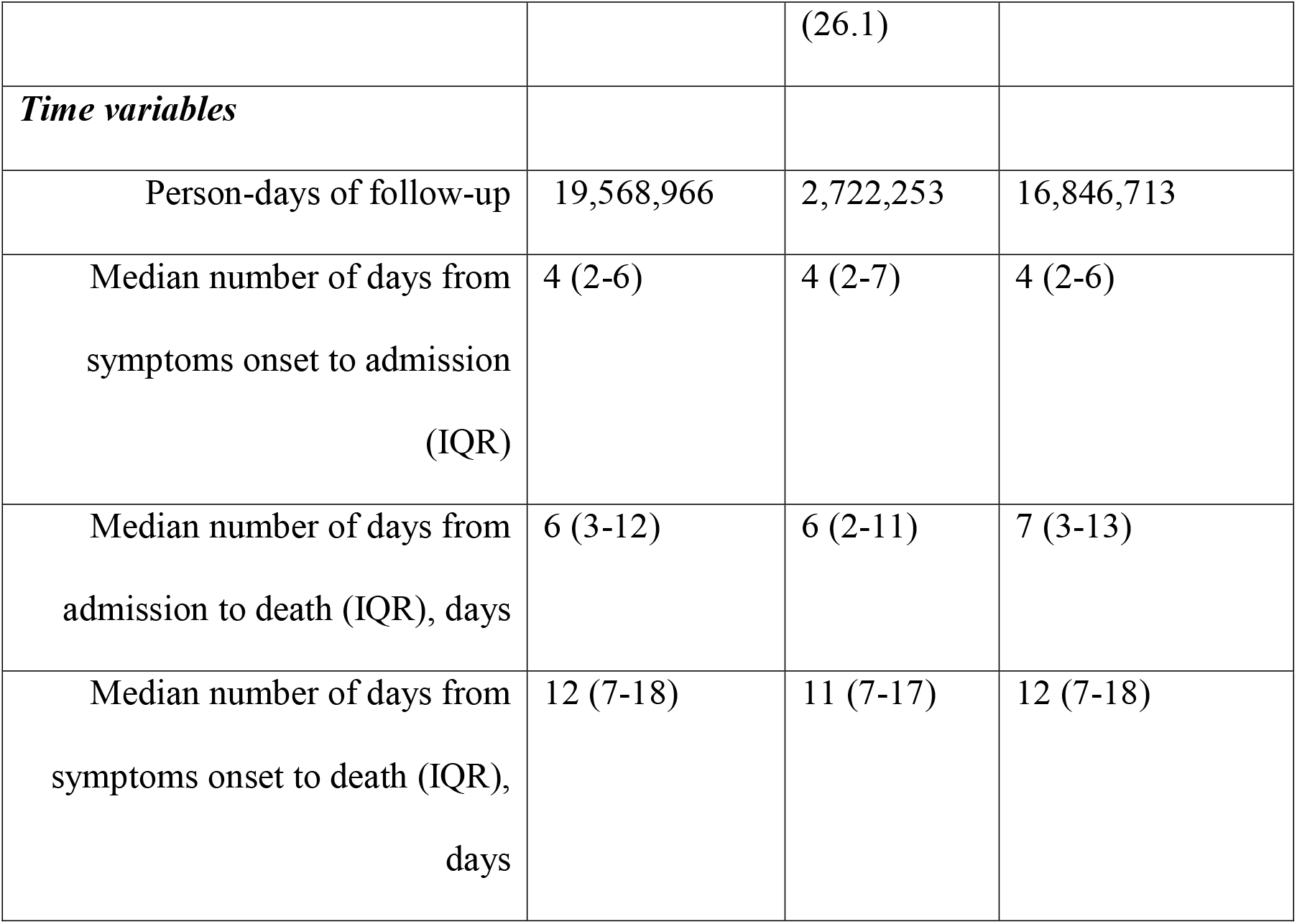
Characteristics of study patients with COVID-19.

In patients without COVID-19, the median age was 40 years (IQR, 30-51); 88,235 (10.0%) had diabetes (Table S2). The proportion of patients with diabetes was 7.6% among outpatients (791,899) and 32.2% among inpatients (86,941).

### Association of diabetes with mortality

As of November 4, 2020, 80,616 deaths occurred among patients with COVID-19 who were followed up for 28 days (19,568,966 person-days of observation); 31,389 (38.9%) had diabetes. During the same period, 20,134 deaths occurred among patients without COVID-19 (24,172,062 person-days of observation); 7,923 (39.4%) had diabetes (Table S3).

In patients with COVID-19, the incidence rate of death was 1,153.1 cases per 100,000 person-days in those with diabetes and 292.2 cases per 100,000 person-days in those without diabetes. In outpatients with COVID-19, the incidence rate of death in those with and without diabetes was, respectively, 194.1 and 39.2 cases per 100,000 person-days. In hospitalized patients with COVID-19, the incidence rate of death in those with and without diabetes was, respectively, 2,552.8 and 1,735.3 cases per 100,000 person-days. In patients without COVID-19, the incidence rate of death was 344.9 cases per 100,000 person-days in those with diabetes and 55.8 cases per 100,000 person-days in those without diabetes.

Among COVID-19 cases, our adjusted Cox proportional-hazards regression analysis showed that patients with diabetes had a 49% higher relative risk of death than those without diabetes (hazard ratio: 1.49 (95% confidence interval [CI], 1.47-1.52) (Table 2). The association of diabetes with mortality was mediated by age, sex, and the type of patient care (outpatient vs. inpatient) (P<0.001 for all interactions). Men were at higher risk of death than women (hazard ratio: 1.65; 95% CI, 1.63-1.68). Compared with subjects 50 to 59 years of age, those 70 to 79 years of age and those 80 years of age or older had 3-fold and 4-fold higher risk of death, respectively (Table S4). A slightly stronger association between diabetes and mortality was noted in women (hazard ratio: 1.64; 95% CI, 1.59-1.68) than in men (hazard ratio: 1.41; 95% CI, 1.38-1.44). We observed a stronger association between diabetes and mortality in outpatients (hazard ratio: 1.66; 95% CI, 1.58-1.74) compared with that in hospitalized patients (hazard ratio: 1.14; 95% CI, 1.12-1.16) (Table 2). Diabetes was associated with lower survival probability in outpatients and inpatients, both in women and men (Figure 1).

**Table 2.**
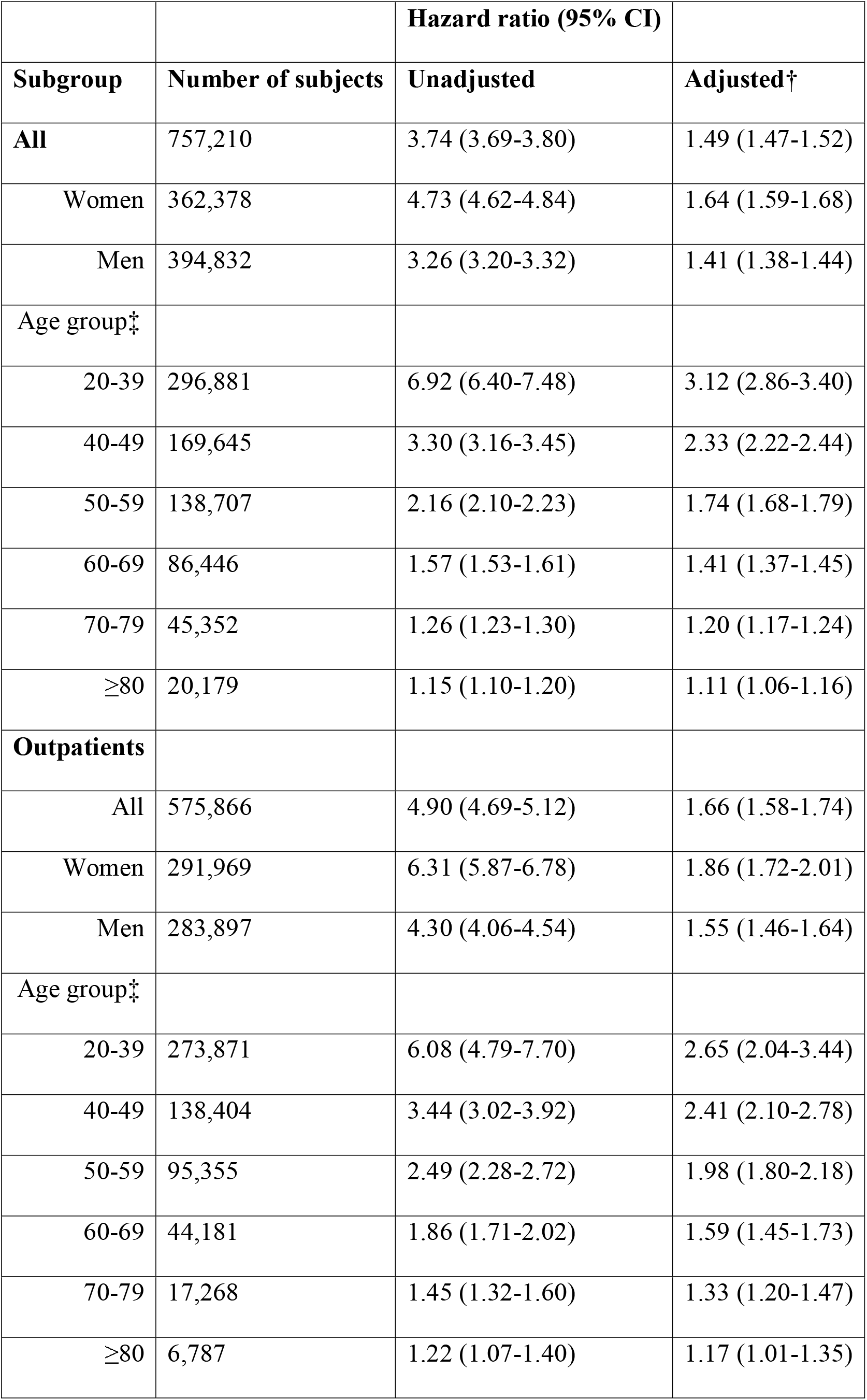
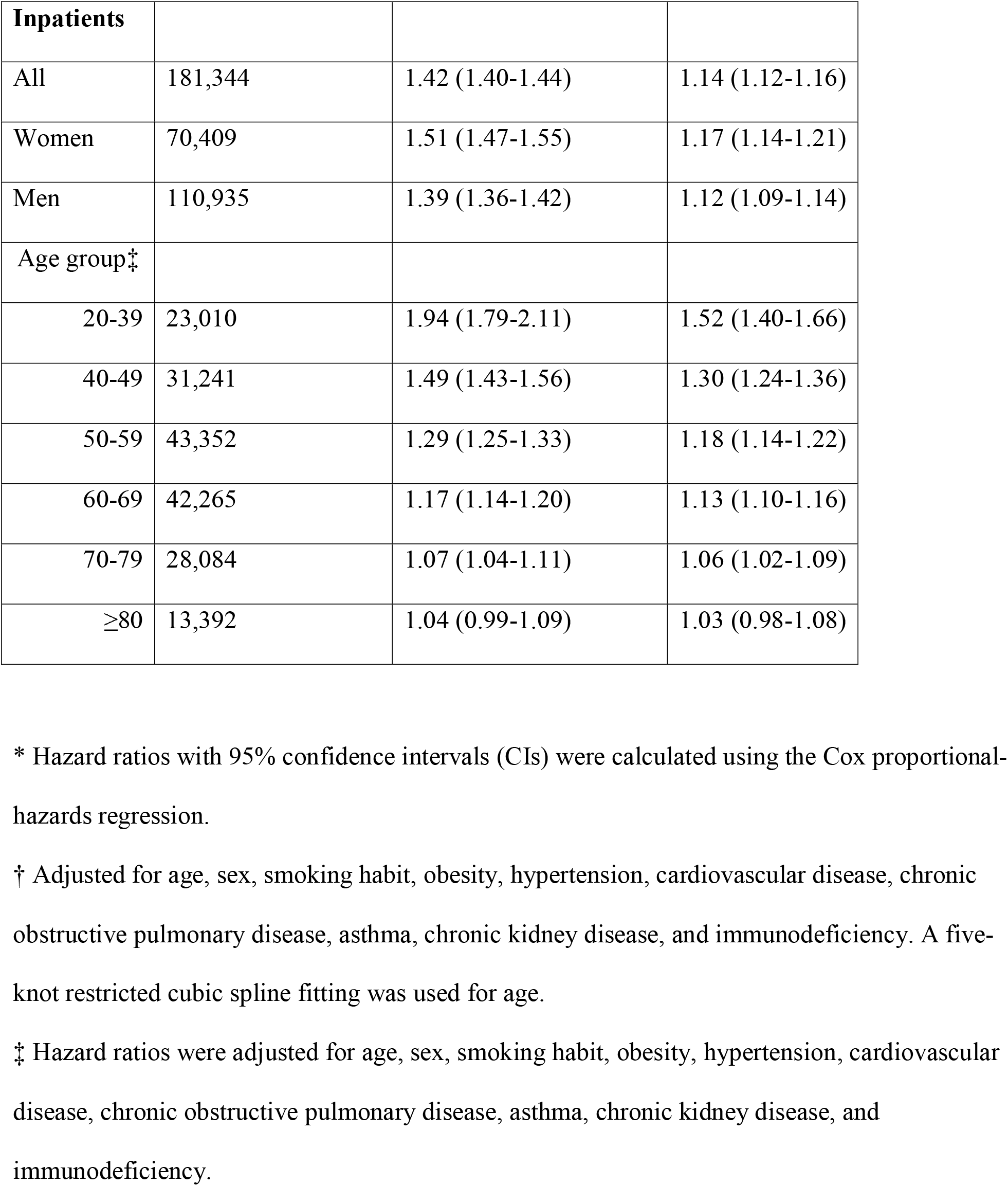
Association of diabetes with mortality among subjects with COVID-19. *.

**Figure 1.**
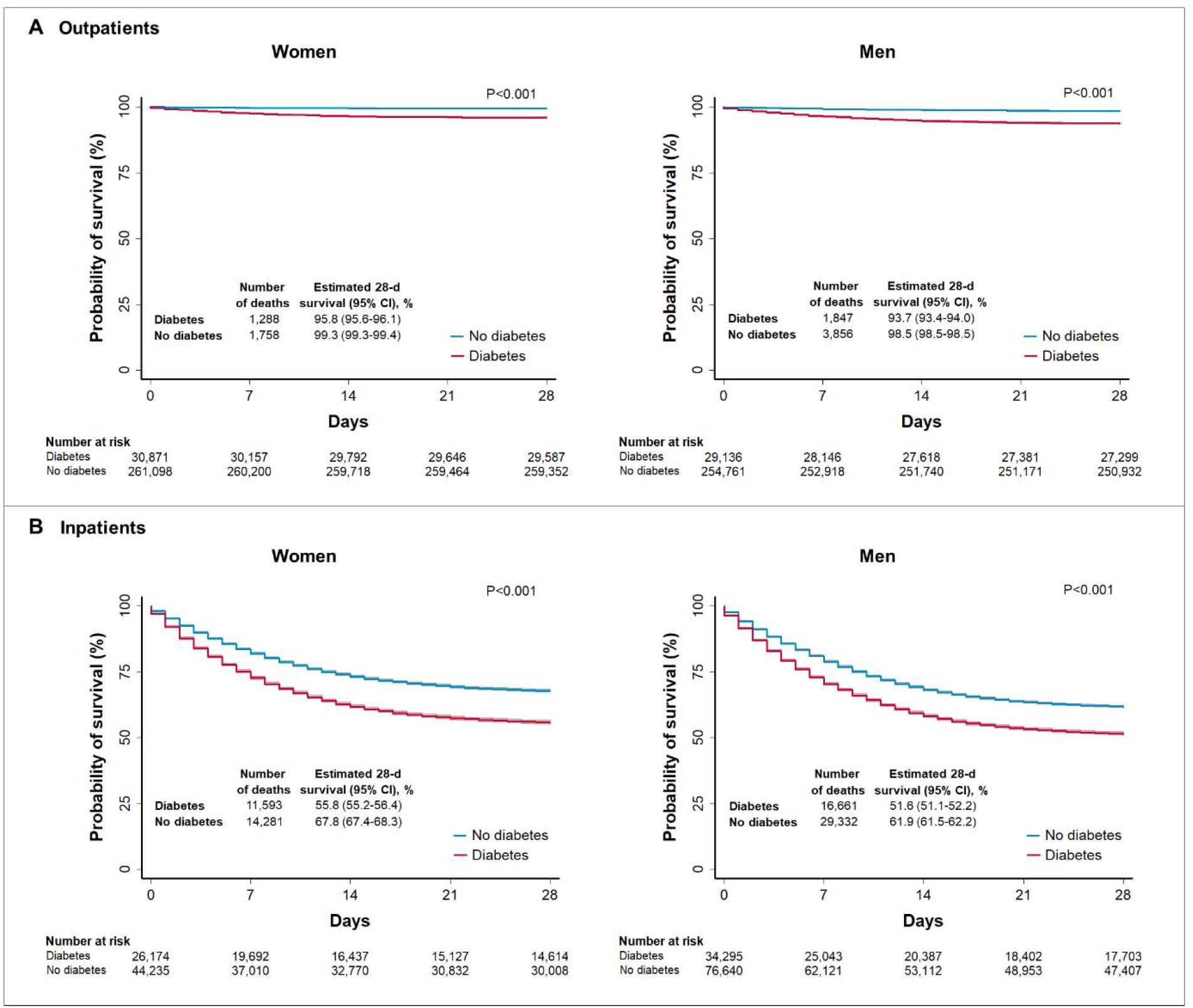
Unadjusted Kaplan-Meier survival curves among outpatients and inpatients with COVID-19 according to sex. Panels show the probability of survival according to sex among adult patients with and without diabetes who had COVID-19. Subjects were admitted from January 1 through October 7, 2020, and followed up for 28 days unless the event (death) occurred first. The solid lines represent survival probabilities and the shaded area represent the 95% confidence intervals (CIs).

In non-COVID-19 cases, the adjusted hazard ratio for mortality was 1.78 (95% CI, 1.73-1.84). This association was also stronger in outpatients (hazard ratio: 1.91; 95% CI, 1.68-2.18) compared with that in hospitalized patients (hazard ratio: 1.11; 95% CI, 1.07-1.14). The 28-day survival for inpatients with diabetes who had COVID-19 was lower (53.4%) compared with that for those without COVID-19 (73.4%) (Figure 2).

**Figure 2.**
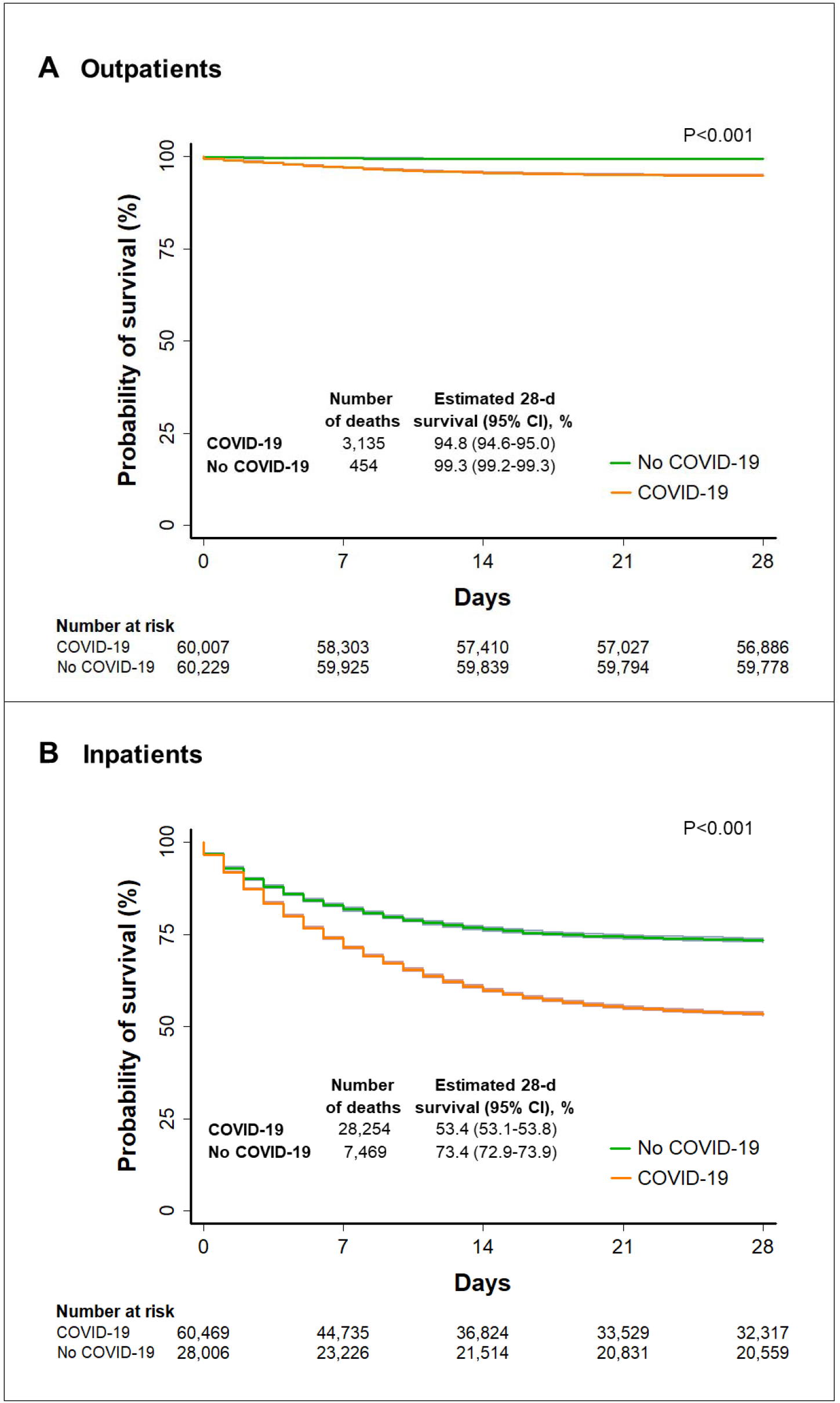
Comparison of unadjusted Kaplan-Meier survival curves among subjects with and without COVID-19. Panels show the probability of survival among outpatients (A) and inpatients (B) who had diabetes with and without COVID-19. Subjects were admitted from January 1 through October 7, 2020, and followed up for 28 days unless the event (death) occurred first. The solid lines represent survival probabilities and the shaded area represent the 95% confidence intervals (CIs).

### Sensitivity and subgroup analyses

In COVID-19 cases, the association of diabetes with mortality persisted after further adjustment for pneumonia, admission to intensive care unit, intubation, and time from symptoms onset to admission (Table S5). Accounting for geographical location did not substantially affect our estimates of the risk of death for outpatients (hazard ratio: 1.62, 95% CI, 1.54-1.70) or inpatients (hazard ratio: 1.13, 95% CI, 1.12-1.15) with COVID-19. The updated guidelines to define suspected cases of viral respiratory disease did not have a substantial effect on the hazard ratios estimates for outpatients (before: 1.66, 95% CI, 1.57-1.74; after: 1.63, 95% CI, 1.42-1.87) or inpatients (before: 1.14, 95% CI, 1.12-1.16; after: 1.15, 95% CI, 1.10-1.19).

In stratified analysis among COVID-19 cases according to sex and age, diabetes was associated with higher mortality in all age groups, among women and men. We observed that the relative risk of death associated with diabetes decreased with age (Table 2) (trend test: P=0.004). These trends were observed in women (P=0.006) and men (P=0.007) (Table S6). Although the relative risk of death decreased with age, the incidence rates of death were higher in older subjects (Table S7). The relative risk of death associated with diabetes decreased with age in outpatients (trend test: P=0.001) and in hospitalized patients (trend test: P=0.006) (Table S8).

In hospitalized patients with COVID-19, the probability of survival at 28 days of follow-up for those with diabetes compared with that among those without diabetes decreased as age increased. We did not observe substantial differences in survival between patients 70 years of age or older with diabetes and those without diabetes (Figure 3). The 28-day survival for inpatients with and without diabetes was, respectively, 73.5% and 85.2% for patients 20-39 years of age; 66.6% and 75.9% for patients 40-49 years of age; 59.4% and 66.5% for patients 50-59 years of age; 50.1% and 54.6% for patients 60-69 years of age; 42.7% and 44.6% for patients 70-79 years of age; and 38.4% and 39.0% for patients 80 years of age or older. In hospitalized patients 60 years of age or older without COVID-19, we did not observe substantial differences in survival between patients with diabetes and those without diabetes (Figure S2).

**Figure 3.**
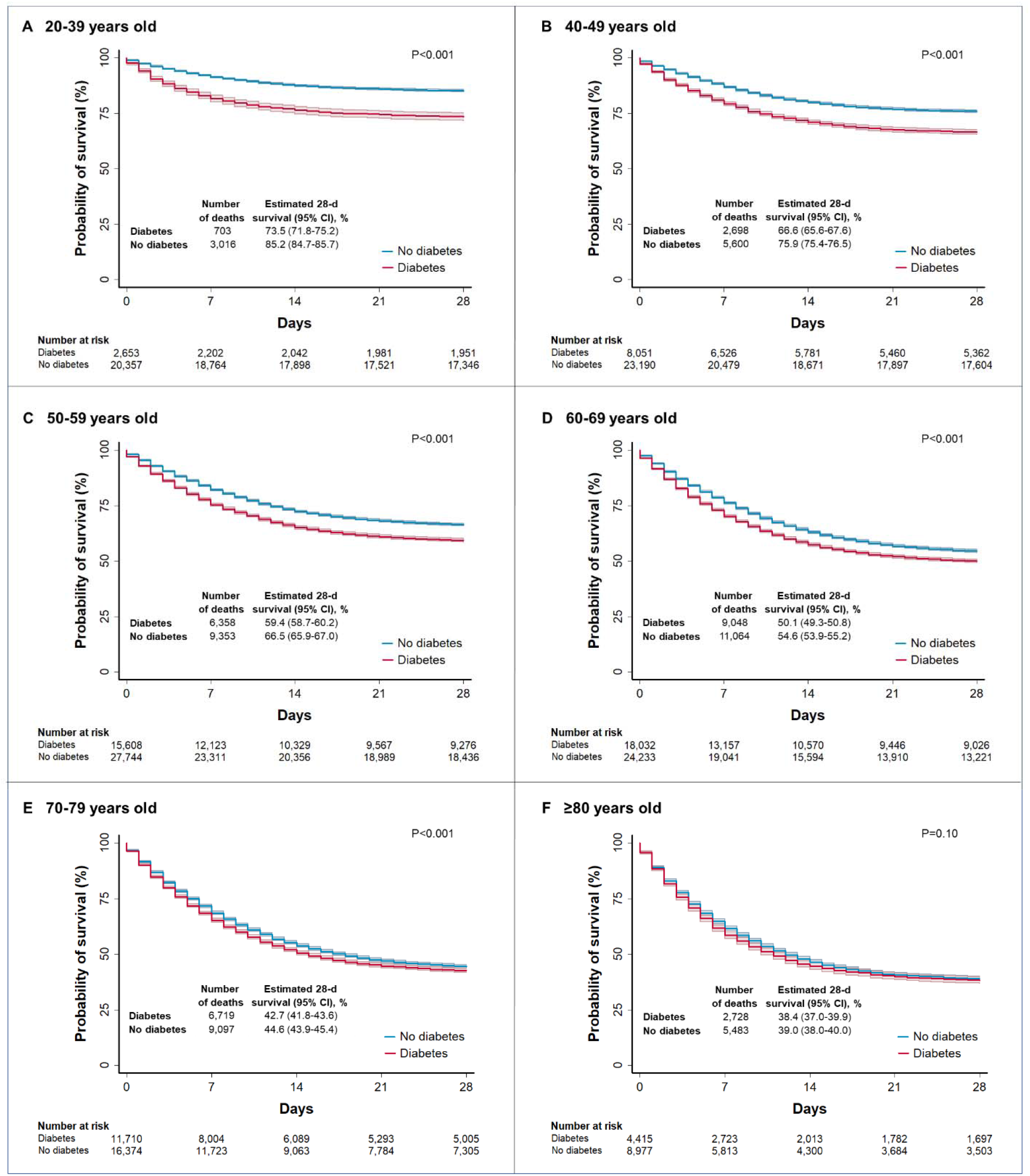
Unadjusted Kaplan-Meier survival curves according to age for inpatients with COVID-19. Panels A to F show the probability of survival stratified according to age groups among adult inpatients with and without diabetes who had COVID-19. Subjects were admitted from January 1 through October 7, 2020, and followed up for 28 days unless the event (death) occurred first. The solid lines represent survival probabilities and the shaded area represent the 95% confidence intervals (CIs).

## DISCUSSION

Our analysis of a large population of symptomatic adult patients with COVID-19 in Mexico (>750,000) shows that those with diabetes have increased risk of death during a follow-up of 28 days. The relative risk of death associated with diabetes was stronger in outpatients than in hospitalized patients, and decreased with age. However, the incidence rate of death was much higher in hospitalized patients than in outpatients, and increased with age.

Although the 28-day survival for hospitalized patients with diabetes in the youngest group (20 to 39 years) was about 12 percentage points lower compared with that for those without diabetes, the survival difference between patients with diabetes and those without diabetes was less than 2 percentage points in those 70 years of age or older (Figure 3). Lower hazard ratios for death associated with diabetes have been reported among older patients with COVID-19 in England ^30^. Our study shows a detailed comparison of the association of diabetes with mortality across age groups in a Hispanic-Latino population in Mexico, a country that has one of the highest numbers of deaths in the world due to COVID-19.^31^

Previous studies have shown diabetes is very common in patients with COVID-19.^26,32^ In our study, the proportion of patients with diabetes among hospitalized patients with COVID-19 was similarly high in those with and without COVID-19 (∼30%). Among deceased patients, the proportion of patients with diabetes was also similar in both groups (∼40%). Although some studies have detected an association between diabetes and mortality in subjects with COVID-19,^4,6,7,9-13^ others have not found a significant association.^2,3,8,15-18^ In our study, diabetes was associated with mortality in patients with COVID-19. However, this association was not stronger than that observed in patients without COVID-19. Our findings also raise concern that the association of diabetes with COVID-19-related mortality varies with age (Table 2). We observed a modest association in patients 70 to 79 years of age. No association was observed in patients 80 years of age or older. The latter age group represented 11% of all COVID-19-related deaths in our study (Table S3). Data from preliminary reports^13,26,33^ suggest that 30-50% of the total number of COVID-19-related deaths occur in patients 80 years of age or older. Thus, our findings may help guide decisions when assessing the risk of death among patients with COVID-19 who have diabetes.

The present study has many strengths. The high number of fatal cases among patients with COVID-19 (>80,000 deaths) and the high number of patients who had diabetes (>120,000) in the population studied enabled us to conduct a stratified analysis according to age groups to obtain precise estimates of the association of diabetes with mortality. The association of diabetes with mortality across age groups in Latino populations has remained unknown. Our stratified analysis by age was performed in a population with a number of deaths that was 15 times higher than the number analyzed in a previous study conducted in a predominantly Caucasian population.^30^ We also performed stratified analysis among outpatients and inpatients. We observed a substantial difference in the magnitude of the association of diabetes with mortality in outpatients (66% higher risk) and inpatients (14% higher risk) with COVID-19, suggesting this association is weaker in patients with severe COVID-19.

Limitations of this study include self-reported diabetes, unknown type of diabetes, and unknown diabetes status. The proportion of patients with diabetes in the overall population (with and without COVID-19) was 12.8%, slightly lower than the adult prevalence of diabetes in Mexico (15.2%).^34^ Our regression models were not adjusted for ethnicity or clinical and laboratory variables since data were not available. Proper blood glucose control has been associated with lower COVID-19-related mortality.^35^ Another limitation of our study is that we cannot exclude the possibility that the number of deaths in patients who had COVID-19 could be underreported. Finally, since our analysis was restricted to patients who presented symptoms for suspected viral respiratory disease and only 10% of patients with mild symptoms of viral respiratory disease were reported to the surveillance system, our findings may not be generalizable to populations with asymptomatic or mild COVID-19.

## CONCLUSION

In symptomatic adult patients with COVID-19 in Mexico, diabetes was associated with higher mortality. The relative risk of death associated with diabetes decreased with age.

## Data Availability

Data are publicly available from the Secretary of Health of Mexico.

https://www.gob.mx/salud/documentos/datos-abiertos-bases-historicas-direccion-general-de-epidemiologia

## ACKNOWLEDGMENTS

We thank the Secretary of Health of the Government of Mexico for providing free access to data on suspected cases of viral respiratory disease in Mexico.

## FUNDING

No funding was received for this study.

## AUTHORSHIP CONFIRMATION STATEMENT

OOW: study design, data collection, statistical analyses, data interpretation, final draft writing. JPCB: contributed with study design and data interpretation. OOW and JPCB have reviewed and approved the manuscript prior to submission.

## AUTHOR DISCLOSURE STATEMENTS

Authors have nothing to disclose.

## Notes

### Competing Interest Statement

The authors have declared no competing interest.

### Author Declarations

Since this study involved the analysis of publicly available de-identified data only, institutional-review-board review was not required, as outlined in the Federal Policy for the Protection of Human Subjects (detailed in 45 CFR part 46).

