## Supplementary appendix for "Diabetes and Mortality Among 1.6 Million Adult Patients Screened for SARS-CoV-2 in Mexico"

Table of Contents

Figure S1. Flow diagram of selection of study patients. 3

Figure S2. Unadjusted Kaplan-Meier survival curves according to age among inpatients without COVID-19. 4

Table S1. Characteristics of study patients eligible for analysis. 5

Table S2. Characteristics of study patients without COVID-19. 6

Table S3. Characteristics of deceased subjects. 7

Table S4. Association of age, sex, pre-existing clinical conditions, and smoking 8

Table S5. Sensitivity analysis for the association of diabetes with mortality among subjects with COVID-19. 9

Table S7. Incidence rates of mortality among subjects with COVID-19 according to age and sex. 11

Table S8. Association of diabetes with mortality among outpatients and inpatients with COVID-19 according to age. 12

**Figure S1. Flow diagram of selection of study patients.**

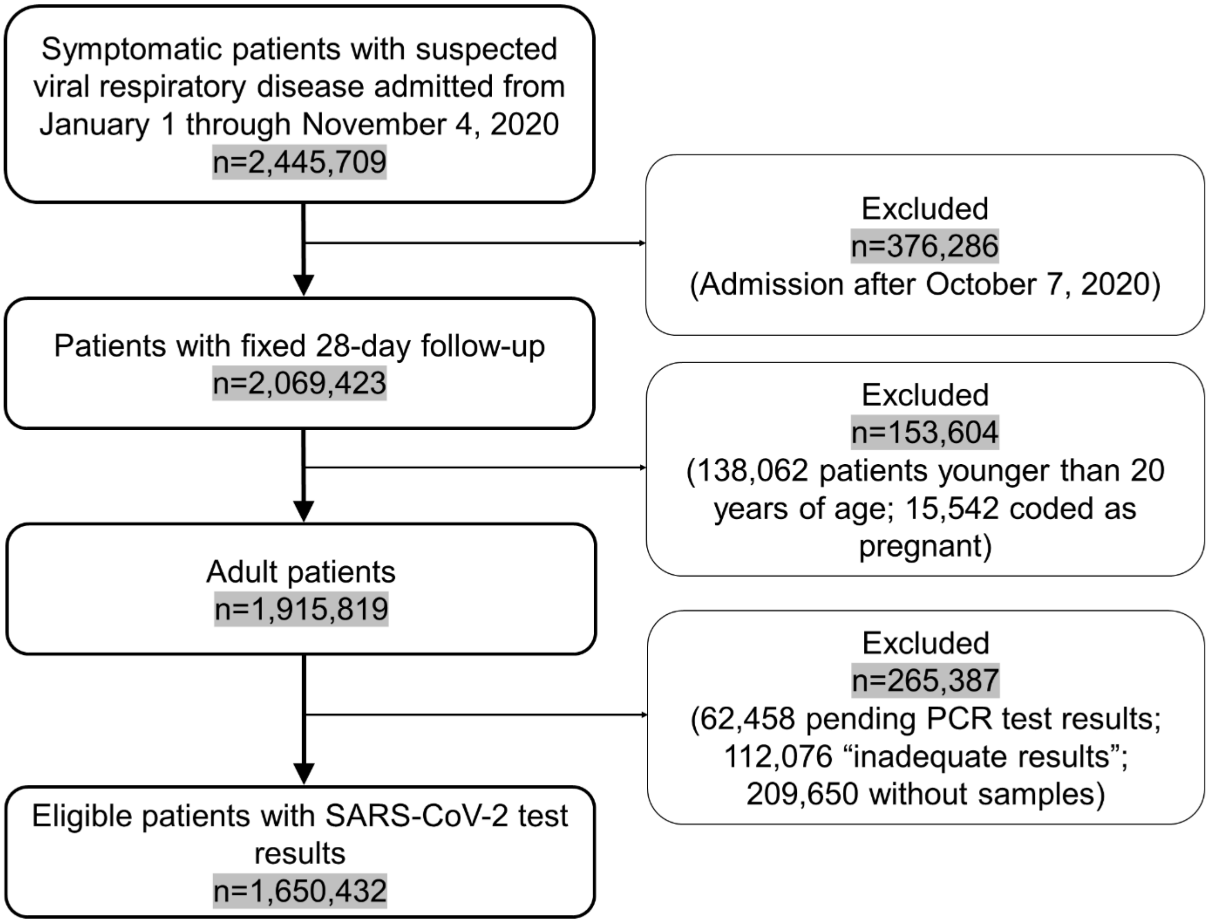

**Figure S2. Unadjusted Kaplan-Meier survival curves according to age among inpatients without COVID-19.** Subjects were enrolled from January 1 through October 7, 2020, and followed up for 28 days unless the event (death) occurred first. The solid lines represent survival probabilities and the shaded area represent the 95% confidence intervals (CIs).

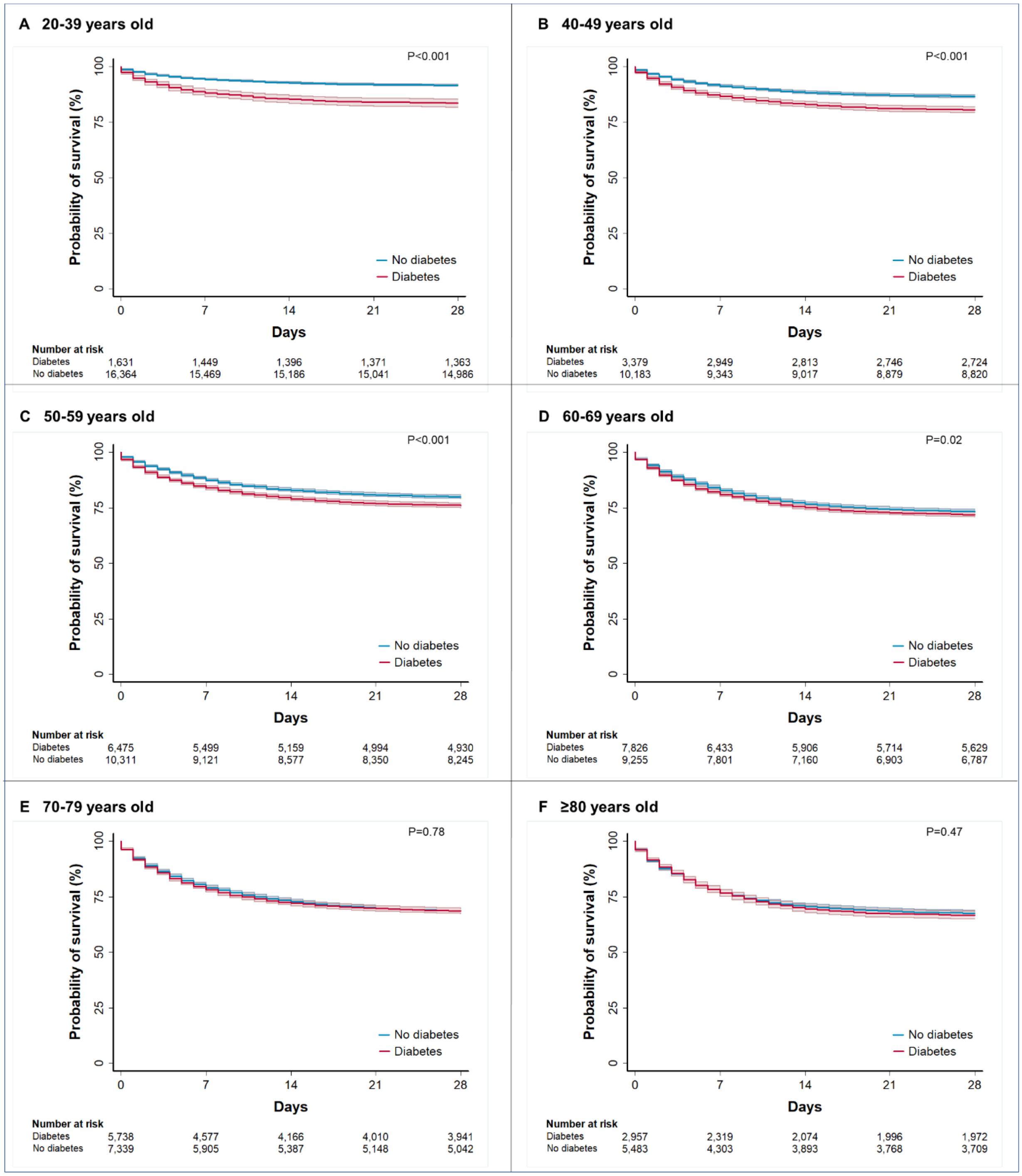

**Table S1.** Characteristics of study patients eligible for analysis.

| Characteristic | Included (n=1,636,050) | Excluded* | | |
|---|---|---|---|---|
| | | All (n=14,185) | With COVID-19 (n=4,461) | Without COVID-19 (n=9,724) |
| Median age (IQR), years | 42 (32-54) | 48 (35-62) | 51 (39-63) | 46 (33-61) |
| Age distribution, n (%) | | | | |
| 20-39 years | 727,106 (44.4) | 4,844 (34.2) | 1,183 (26.5) | 3,661 (37.7) |
| 40-49 years | 365,495 (22.3) | 2,738 (19.3) | 886 (19.9) | 1,852 (19.1) |
| 50-59 years | 275,110 (16.8) | 2,601 (18.3) | 975 (21.9) | 1,626 (16.7) |
| 60-69 years | 154,676 (9.5) | 1,857 (13.1) | 737 (16.5) | 1,120 (11.5) |
| 70-79 years | 77,195 (4.7) | 1,200 (8.5) | 454 (10.2) | 746 (7.7) |
| ≥80 years | 36,468 (2.2) | 945 (6.7) | 226 (5.1) | 719 (7.4) |
| Male sex, n (%) | 802,793 (49.1) | 7,269 (51.2) | 2,592 (58.1) | 4,677 (48.1) |
| ***Outcomes, n (%)*** | | | | |
| SARS-CoV-2 infection | 757,210 (46.3) | 4,461 (31.5) | 4,461 (100) | -- |
| Outpatient | 1,367,765 (83.6) | 7,602 (53.6) | 2,475 (55.5) | 5,127 (52.7) |
| Hospitalized | 268,285 (16.4) | 6,583 (46.4) | 1,986 (44.5) | 4,597 (47.3) |
| Died | 100,750 (6.2) | 2,150 (15.2) | 1,250 (28.0) | 900 (9.3) |
| ***Time variables*** | | | | |
| Median number of days from symptoms onset to admission (IQR) | 3 (1-5) | 2 (1-5) | 4 (2-6) | 2 (1-4) |
| Median number of days from admission to death (IQR), days | 6 (2-12) | 5 (2-11) | 6 (2-12) | 4 (2-10) |
| Median number of days from symptoms onset to death (IQR), days | 11 (7-17) | 10 (5-16) | 11 (7-18) | 7 (4-14) |

* Patients with missing data on predictors included in our Cox proportional-hazard regression models. Missing data were not imputed: diabetes (8,216; 0.34%), smoking habit (7,939; 0.32%), obesity (7,349; 0.30%), hypertension (7,704; 0.32%), cardiovascular disease (7,600; 0.31%), chronic obstructive pulmonary disease (7,607; 0.31%), asthma (7,525; 0.31%), chronic kidney disease (7,487; 0.31%), immunodeficiency (7,968; 0.33%), pneumonia (18,319; 0.75%), intubation (6,695; 0.27%), and admission to intensive care unit (7,254; 0.30%).

**Table S2.** Characteristics of study patients without COVID-19.

| Characteristic | All (n=878,840) | With diabetes (n=88,235) | Without diabetes (n=790,605) |
|---|---|---|---|
| Median age (IQR), years | 40 (30-51) | 56 (47-66) | 38 (30-49) |
| Age distribution, n (%) | | | |
| 20-39 years | 430,225 (49.0) | 9,126 (10.3) | 421,099 (53.3) |
| 40-49 years | 195,850 (22.3) | 17,577 (19.9) | 178,273 (22.6) |
| 50-59 years | 136,403 (15.5) | 25,127 (28.5) | 111,276 (14.1) |
| 60-69 years | 68,230 (7.8) | 20,388 (23.1) | 47,842 (6.1) |
| 70-79 years | 31,843 (3.6) | 11,188 (12.7) | 20,655 (2.6) |
| ≥80 years | 16,289 (1.9) | 4,829 (5.5) | 11,460 (1.5) |
| Male sex, n (%) | 407,961 (46.4) | 41,674 (47.2) | 366,287 (46.3) |
| Smoking habit, n (%) | 85,816 (9.8) | 9,069 (10.3) | 76,747 (9.7) |
| Pneumonia, n (%) | 60,618 (6.9) | 18,184 (20.6) | 42,434 (5.4) |
| ***Pre-existing comorbidities, n (%)*** | | | |
| Diabetes | 88,235 (10.0) | -- | -- |
| Obesity | 122,550 (13.9) | 22,242 (25.2) | 100,308 (12.7) |
| Hypertension | 125,014 (14.2) | 46,801 (53.0) | 78,213 (9.9) |
| Cardiovascular disease | 16,467 (1.9) | 5,863 (6.6) | 10,604 (1.3) |
| Chronic kidney disease | 14,804 (1.7) | 7,538 (8.5) | 7,266 (0.9) |
| COPD | 10,998 (1.3) | 4,076 (4.6) | 6,922 (0.9) |
| Asthma | 26,515 (3.0) | 2,938 (3.3) | 23,577 (3.0) |
| Immunodeficiency | 10,401 (1.2) | 2,646 (3.0) | 7,755 (1.0) |
| Any comorbidity including diabetes | 287,000 (32.7) | -- | -- |
| Any comorbidity excluding diabetes | 258,925 (29.5) | 60,160 (68.2) | 198,765 (25.1) |
| ***Outcomes, n (%)*** | | | |
| Outpatient | 791,899 (90.1) | 60,229 (68.3) | 731,670 (92.6) |
| Hospitalized | 86,941 (9.9) | 28,006 (31.7) | 58,935 (7.5) |
| Admitted to intensive care unit | 5,957 (0.7) | 2,021 (2.3) | 3,936 (0.5) |
| Intubated | 8,305 (0.9) | 2,886 (3.3) | 5,419 (0.7) |
| Died | 20,134 (2.3) | 7,923 (9.0) | 12,211 (1.5) |
| ***Time variables*** | | | |
| Person-days of follow-up | 24,172,062 | 2,297,161 | 21,874,901 |
| Median number of days from symptoms onset to admission (IQR) | 3 (1-5) | 3 (1-5) | 3 (1-5) |
| Median number of days from admission to death (IQR), days | 5 (2-11) | 4 (1-10) | 5 (2-11) |
| Median number of days from symptoms onset to death (IQR), days | 9 (5-15) | 9 (5-15) | 9 (5-16) |

COPD, chronic obstructive pulmonary disease.

**Table S3.** Characteristics of deceased subjects.

| Characteristic | With COVID-19 (n=80,616) | Without COVID-19 (n=20,134) |
|---|---|---|
| Median age (IQR), years | 63 (54-73) | 64 (53-74) |
| Age distribution, n (%) | | |
| 20-39 years | 4,301 (5.3) | 1,789 (8.9) |
| 40-49 years | 9,409 (11.7) | 2,182 (10.8) |
| 50-59 years | 17,766 (22.0) | 3,868 (19.2) |
| 60-69 years | 22,422 (27.8) | 4,983 (24.8) |
| 70-79 years | 17,532 (21.8) | 4,365 (21.7) |
| ≥80 years | 9,186 (11.4) | 2,947 (14.6) |
| Male sex, n (%) | 51,696 (64.1) | 12,164 (60.4) |
| Smoking habit, n (%) | 6,537 (8.1) | 2,194 (10.9) |
| Pneumonia, n (%) | 59,673 (74.0) | 13,141 (65.3) |
| ***Pre-existing comorbidities, n (%)*** | | |
| Diabetes | 31,389 (38.9) | 7,923 (39.4) |
| Obesity | 19,840 (24.6) | 3,649 (18.1) |
| Hypertension | 36,764 (45.6) | 9,204 (45.7) |
| Cardiovascular disease | 4,347 (5.4) | 1,710 (8.5) |
| Chronic kidney disease | 5,868 (7.3) | 2,514 (12.5) |
| COPD | 3,966 (4.9) | 1,577 (7.8) |
| Asthma | 1,610 (2.0) | 393 (2.0) |
| Immunodeficiency | 1,968 (2.4) | 1,110 (5.5) |
| Any comorbidity including diabetes | 58,180 (72.2) | 14,747 (73.2) |
| Any comorbidity excluding diabetes | 51,101 (63.4) | 13,244 (65.8) |
| ***Outcomes, n (%)*** | | |
| Outpatient | 8,749 (10.9) | 1,279 (6.4) |
| Hospitalized | 71,867 (89.2) | 18,855 (93.7) |
| Admitted to intensive care unit | 7,977 (9.9) | 1,927 (9.6) |
| Intubated | 25,335 (31.4) | 5,019 (24.9) |
| ***Time variables*** | | |
| Median number of days from symptoms onset to admission (IQR) | 4 (2-7) | 3 (1-6) |
| Median number of days from admission to death (IQR), days | 6 (3-11) | 4 (1-10) |
| Median number of days from symptoms onset to death (IQR), days | 11 (7-17) | 9 (5-15) |

COPD, chronic obstructive pulmonary disease.

**Table S4.** Association of age, sex, pre-existing clinical conditions, and smoking habit with mortality among subjects with COVID-19.*

| | Hazard ratio (95% CI) | |
|---|---|---|
| **Variable** | **Unadjusted** | **Adjusted†** |
| Age group, yr | | |
| 20-39 | 0.11 (0.10-0.11) | 0.13 (0.13-0.14) |
| 40-49 | 0.42 (0.41-0.43) | 0.46 (0.45-0.47) |
| 50-59 | Reference | Reference |
| 60-69 | 2.17 (2.13-2.22) | 1.97 (1.93-2.01) |
| 70-79 | 3.51 (3.44-3.59) | 3.05 (2.99-3.12) |
| ≥80 | 4.42 (4.31-4.53) | 4.02 (3.91-4.12) |
| Male sex | 1.68 (1.66-1.71) | 1.65 (1.63-1.68) |
| Diabetes | 3.74 (3.69-3.80) | 1.49 (1.47-1.52) |
| Obesity | 1.49 (1.46-1.51) | 1.39 (1.37-1.41) |
| Hypertension | 3.68 (3.63-3.73) | 1.23 (1.21-1.25) |
| Cardiovascular disease | 3.13 (3.04-3.23) | 0.99 (0.96-1.02) |
| Chronic kidney disease | 4.81 (4.68-4.94) | 2.01 (1.95-2.06) |
| Chronic obstructive pulmonary disease | 3.98 (3.85-4.11) | 1.21 (1.17-1.25) |
| Asthma | 0.77 (0.73-0.81) | 0.86 (0.82-0.91) |
| Immunodeficiency | 2.55 (2.44-2.67) | 1.39 (1.33-1.45) |
| Smoking habit | 1.08 (1.05-1.11) | 0.97 (0.94-0.99) |

* Hazard ratios with 95% confidence intervals (CIs) were calculated using the Cox proportional-hazards regression.

† Hazard ratios were adjusted for all variables included in the model and age (a five-knot restricted cubic spline fitting was used for age), except for age group.

**Table S5.** Sensitivity analysis for the association of diabetes with mortality among subjects with COVID-19.

| | **Adjusted hazard ratio (95% CI) *** |
|---|---|
| Full model † | 1.49 (1.47-1.52) |
| Full model adjusted for pneumonia | 1.26 (1.25-1.28) |
| Full model adjusted for admission to intensive care unit | 1.47 (1.45-1.49) |
| Full model adjusted for intubation | 1.42 (1.40-1.44) |
| Full model adjusted for time from symptoms onset to admission | 1.49 (1.47-1.52) |

* Hazard ratios with 95% confidence intervals (CIs) were calculated using the Cox proportional-hazards regression.

† Full model denotes the association of diabetes with mortality adjusting for age, sex, smoking habit, obesity, hypertension, cardiovascular disease, chronic obstructive pulmonary disease, asthma, chronic kidney disease, and immunodeficiency. A five-knot restricted cubic spline fitting was used for age.

**Table S6.** Association of diabetes with mortality among subjects with COVID-19 according to age and sex. *

| | **Women** | | **Men** | |
|---|---|---|---|---|
| | **Number of subjects** | **Adjusted hazard ratio (95% CI)** | **Number of subjects** | **Adjusted hazard ratio (95% CI)** |
| **Age group, yr** | | | | |
| 20-39 | 146,521 | 4.09 (3.54-4.74) | 150,360 | 2.65 (2.38-2.96) |
| 40-49 | 82,251 | 2.91 (2.68-3.16) | 87,394 | 2.09 (1.97-2.21) |
| 50-59 | 65,587 | 2.06 (1.95-2.18) | 73,120 | 1.58 (1.52-1.65) |
| 60-69 | 39,173 | 1.53 (1.46-1.61) | 47,273 | 1.33 (1.29-1.38) |
| 70-79 | 19,749 | 1.26 (1.19-1.32) | 25,603 | 1.17 (1.12-1.22) |
| ≥80 | 9,097 | 1.12 (1.05-1.20) | 11,082 | 1.10(1.03-1.16) |

* Hazard ratios with 95% confidence intervals (CIs) were calculated using the Cox proportional-hazards regression. Estimates within each age group were adjusted for age, sex, smoking habit, obesity, hypertension, cardiovascular disease, chronic obstructive pulmonary disease, asthma, chronic kidney disease, and immunodeficiency.

**Table S7.** Incidence rates of mortality among subjects with COVID-19 according to age and sex.

| | Incidence rate per 100,000 person-years * | | | | | |
|---|---|---|---|---|---|---|
| | All | | Women | | Men | |
| | With diabetes | Without diabetes | With diabetes | Without diabetes | With diabetes | Without diabetes |
| **Age group, yr** | | | | | | |
| 20-39 | 311.0 | 44.1 | 263.8 | 25.6 | 355.7 | 62.4 |
| 40-49 | 541.0 | 159.9 | 421.8 | 91.8 | 648.3 | 226.6 |
| 50-59 | 872.3 | 392.2 | 696.6 | 242.4 | 1,041.0 | 533.6 |
| 60-69 | 1,523.3 | 942.4 | 1,290.3 | 685.6 | 1,750.9 | 1,161.0 |
| 70-79 | 2,248.7 | 1,733.5 | 1,933.4 | 1,395.6 | 2,549.8 | 1,995.6 |
| ≥80 | 2,773.6 | 2,360.6 | 2,380.2 | 1,927.4 | 3,197.1 | 2,730.0 |

* Incidence rates were estimated for subjects with PCR-confirmed COVID-19 enrolled from January 1 through October 7, 2020, who were followed up for 28 days unless the event (death) occurred first.

**Table S8.** Association of diabetes with mortality among outpatients and inpatients with COVID-19 according to age. *

| | | Hazard ratio (95% CI) | |
|---|---|---|---|
| **Subgroup** | **Number of subjects** | **Unadjusted** | **Adjusted†** |
| Outpatients | | | |
| 20-39 | 273,871 | 6.08 (4.79-7.70) | 2.65 (2.04-3.44) |
| 40-49 | 138,404 | 3.44 (3.02-3.92) | 2.41 (2.10-2.78) |
| 50-59 | 95,355 | 2.49 (2.28-2.72) | 1.98 (1.80-2.18) |
| 60-69 | 44,181 | 1.86 (1.71-2.02) | 1.59 (1.45-1.73) |
| 70-79 | 17,268 | 1.45 (1.32-1.60) | 1.33 (1.20-1.47) |
| ≥80 | 6,787 | 1.22 (1.07-1.40) | 1.17 (1.01-1.35) |
| Inpatients | | | |
| 20-39 | 23,010 | 1.94 (1.79-2.11) | 1.52 (1.40-1.66) |
| 40-49 | 31,241 | 1.49 (1.43-1.56) | 1.30 (1.24-1.36) |
| 50-59 | 43,352 | 1.29 (1.25-1.33) | 1.18 (1.14-1.22) |
| 60-69 | 42,265 | 1.17 (1.14-1.20) | 1.13 (1.10-1.16) |
| 70-79 | 28,084 | 1.07 (1.04-1.11) | 1.06 (1.02-1.09) |
| ≥80 | 13,392 | 1.04 (0.99-1.09) | 1.03 (0.98-1.08) |

* Hazard ratios with 95% confidence intervals (CIs) were calculated using the Cox proportional-hazards regression.

† Estimates within each age group were adjusted for age, sex, smoking habit, obesity, hypertension, cardiovascular disease, chronic obstructive pulmonary disease, asthma, chronic kidney disease, and immunodeficiency.
